# Effect of smoking on drug-resistant tuberculosis treatment outcomes and potential mechanistic pathways: A multicountry cohort study

**DOI:** 10.1101/2025.08.20.25334077

**Authors:** Matthew L. Romo, Allison LaHood, Helen R. Stagg, Carole D. Mitnick, Letizia Trevisi, Cathy Hewison, Shrivani Padayachee, Edwin Herrera Flores, Lawrence Oyewusi, Palwasha Y. Khan, Helena Huerga, Mathieu Bastard, Michael L. Rich, Girum Bayissa Tefera, Mahmud Rashitov, Ohanna Kirakosyan, Aga Krisnanda, Atyrkul Toktogonova, Muhammad Rafi Siddiqui, Camilo Gómez-Restrepo, Tina Kotrikadze, Molly F. Franke, the endTB Observational Study Team

## Abstract

**Background:** People who smoke are at increased risk of unfavorable tuberculosis treatment outcomes compared with those who do not, but the pathways that explain this disparity are unclear.

**Objective:** To estimate the difference in a successful end-of-treatment outcome by smoking status among people with multidrug- or rifampicin-resistant tuberculosis (MDR/RR-TB) and to examine if this difference changes if people who smoked had the same retention in treatment as those who did not smoke.

**Design and methods:** Using data from the prospective endTB Observational Study, we estimated the difference in treatment success by cigarette smoking status, adjusting for baseline confounders including demographics, social history, and comorbidities. To examine how this difference changed if everyone was retained in treatment, we censored participants who were lost to follow-up and applied inverse probability of censoring weights to simulate this scenario.

**Results:** Among 1786 participants in 12 countries, 539 (30.2%) reported smoking at least 1 cigarette daily. People who smoked were more frequently in post-Soviet countries, and had a complex social history (e.g., incarceration and substance use) and infectious comorbidities (e.g., hepatitis C). At the end of treatment, 73.5% of people who smoked and 80.3% of people who did not smoke had treatment success (risk difference in percentage points: -6.8, 95% confidence interval [CI]: -11.1, -2.6). After adjusting for baseline confounders, the risk difference was similar (-5.2 percentage points) but the 95% CI was less precise (-14.1, 3.2). When simulating a scenario in which everyone was retained in treatment, the risk difference was attenuated (-1.9 percentage points; 95% CI: -11.1, 4.7).

**Conclusion:** People who smoked had a lower frequency of MDR/RR-TB treatment success than those who did not smoke. Eliminating loss to follow-up reduced this difference by smoking status, suggesting that pathways related to retention in treatment were a major driver of this disparity.

**What is already known on this topic:** Most observational research supports that cigarette smoking negatively impacts TB treatment outcomes, but it is unclear why.

**What this study adds:** People who smoked had a lower frequency of MDR/RR-TB treatment success than those who did not smoke. Eliminating loss to follow-up attenuated this difference.

**How this study might affect research, practice, or policy:** Implementing interventions that address causes of loss to follow-up, in addition to smoking cessation services, could improve MDR/RR-TB treatment outcomes among people who smoke.

## INTRODUCTION

Tuberculosis (TB) and tobacco smoking are both major contributors to morbidity and mortality.^1,2^ Their intersection is also substantial, with a higher prevalence of tobacco smoking in high TB burden countries and among populations that are disproportionately affected by TB, such as men, people living with HIV, incarcerated people, and mine workers.^1,2^ The dual burden of TB and smoking is not coincidental. Epidemiologic evidence consistently shows that tobacco smoke increases the risk of TB infection and disease.^3,4^ Tobacco smoking has also been associated with more severe TB disease presentation, including greater sputum mycobacterial load, more cavitary lesions, and higher probability of disease requiring hospitalization;^5–7^ and worse TB treatment outcomes, specifically increased risk of delayed smear and culture conversion, an unsuccessful end-of-treatment outcome, and recurrence.^8–11^

The negative effect of smoking on TB treatment outcomes could be explained by direct biological effects of tobacco smoke. Tobacco smoke causes profound structural and functional damage to the respiratory tract^12^ and impairs the pulmonary immune response to TB infection.^13^ These changes favor the pathogen’s survival, persistence, and proliferation, and could compromise TB treatment effectiveness. Another pathway to unfavorable treatment outcomes is greater adherence challenges among people who smoke, culminating in early discontinuation from treatment, i.e., loss to follow-up after treatment initiation.^14^ This pathway is complex and likely related to multiple factors at different levels among people who smoke, rather than caused by smoking itself. Factors involved in this pathway might include competing economic responsibilities, suboptimal social support, negative experiences accessing care, and clinical evolution and adverse events on TB treatment.^15–19^ The observed association between smoking and TB treatment outcomes may also be spurious, as smoking is frequently accompanied by other factors known to affect TB treatment outcomes, such as alcohol and other substance use.^20–22^

Multidrug- or rifampicin-resistant TB (MDR/RR-TB) is more difficult to treat than drug-susceptible TB,^1^ and individuals often endure an extended duration of complicated drug regimens with multiple toxicities.^23^ Impacts may be experienced by the individual as well as by their family and others around them.^24–26^ Therefore, identifying areas for intervention that effectively increase treatment success is of high clinical and public health importance. Most of the evidence for the negative effect of smoking on TB treatment outcomes comes from studies enrolling people with drug-susceptible TB.^8,10,11^ A meta-analysis of observational studies focused on MDR/RR-TB reported that TB treatment outcomes were similar by smoking status; however, most of the studies defined smoking as ever smoking rather than current smoking at the time of treatment initiation and heterogeneity among the studies was very high.^27^ These observational studies among people with drug-susceptible TB or MDR/RR-TB have largely examined the association between smoking and treatment outcomes without an explicit focus on causality and have not applied methods to clarify the contribution of underlying mechanistic pathways.

Using epidemiologic methods rooted in causal inference, we estimate the effect of smoking on MDR/RR-TB end-of-treatment outcomes. To understand how differential loss to follow-up may contribute to worse treatment outcomes among people who smoke, we estimated the difference in treatment success that would be expected if everyone was retained in treatment.

## METHODS

### Setting and participants

We used data from the endTB Observational Study (https://ClinicalTrials.gov Identifier: NCT03259269), a prospective cohort of people with MDR/RR-TB who were treated with longer regimens containing bedaquiline and/or delamanid.^28,29^ Participants were enrolled from April 2015 through September 2018 in 17 countries. The study sites selected for the cohort reflect the heterogeneity of high-burden MDR/RR-TB settings globally. Participants received treatment regimens with drug selection informed by national TB program guidelines and the endTB clinical guide.^29^ The study captured routine clinical data under programmatic conditions, with standardized data collection that was managed centrally. After enrollment, study visits typically occurred at two weeks and then monthly thereafter throughout treatment. Details on study procedures are available elsewhere.^28^ The study protocol was approved by ethics committees of each consortium partner and country. Participants (or a guardian if the individual was a minor, as defined by local legal requirements) provided written informed consent and older minors provided assent. If individuals were treated multiple times during the study, we included only the first treatment regimen for these analyses. For these analyses, we excluded participants who: were treated in the Democratic People’s Republic of Korea; had confirmed rifampicin-susceptible TB; had only extrapulmonary TB; enrolled in the endTB Observational Study more than one month after MDR/RR-TB treatment initiation; were under 15 years of age; were treated at a site with no participants who reported smoking; were treated at a site that did not collect adherence data or had inconsistent reporting;^30^ had an unknown cigarette smoking status; or transferred out or had an unknown end-of-treatment outcome. Rationale for each exclusion criterion is provided in **Supplementary Table 1**. Patients or the public were not involved in the design, conduct, reporting, or dissemination plans of this secondary data analysis.

### Variables

#### Exposure and outcome

Our exposure of interest was smoking status, which was assessed at the enrollment visit based on a yes/no response when participants were asked if they smoke at least one cigarette daily. End-of-treatment outcomes were assigned by the treating clinician based on World Health Organization outcome definitions.^31^ Successful treatment comprised an outcome of cured or completed. Unsuccessful treatment comprised an outcome of death, treatment failure, or lost to follow-up. Lost to follow-up was defined as a treatment interruption for ≥2 consecutive months.

#### Other variables

Demographics (country, sex, age) and social history variables (currently married or living with partner, employed, homeless in past year, ever incarcerated, refugee/displaced person/migrant status, alcohol use, and drug use) were self-reported at enrollment. We categorized countries as a post-Soviet country or other country, in recognition of the higher proportion of participants who smoked in the former.

Comorbidities at enrollment included HIV; hepatitis B virus infection, based on a positive surface antigen; hepatitis C virus infection, based on a positive antibody, PCR, or viral load; diabetes, based on self-reported diagnosis, random plasma glucose >200 mg/dL (11.1 mmol/L), or hemoglobin A1C ≥6.5%; and underweight, defined as a body mass index <18.5 kg/m^2^ using measured height and weight.

TB disease and treatment characteristics at enrollment included the presence of extrapulmonary TB; presence of bilateral disease, fibrosis, and cavitary disease based on chest x-ray; sputum smear positive and grade; sputum culture positivity; known prior treatment with second-line TB drugs; drug susceptibility results categorized as to whether fluoroquinolone resistance was present or not; and baseline TB regimen composition, including the number of likely effective drugs.

Variables assessed during follow-up included indicators of TB disease severity (i.e., sputum smear positivity, sputum culture positivity, and presence of cavitary disease) and TB treatment adherence. Adherence, typically assessed monthly, was computed as the number of days all medications were taken as prescribed divided by the number of days those medications were prescribed.^30^ This proportion was dichotomized as <80% and ≥80% for each time period it was measured. These variables were treated as time-varying, reflecting changes throughout follow-up.

### Statistical analyses

We estimated the difference in the frequency of treatment success by smoking status in an unadjusted model and a model adjusted for baseline confounders using marginal standardization to compute risk differences and ratios. The bias-corrected and accelerated bootstrap with 1000 resamples was used to obtain 95% confidence intervals (CIs). We used the missing indicator method to account for missing data on baseline confounders. Most missing data were for the past incarceration variable (301/1786 [16.9%] missing; **Table 1**), with two sites having >90% missing responses. Because unmeasured, site-level factors likely contributed, we did not expect that other methods of addressing missing data, specifically multiple imputation, to be advantageous. We used directed acyclic graphs (**Figure 1A-D**) to specify the underlying causal structure between smoking and MDR/RR-TB treatment outcome and identified potential baseline demographic, social, and clinical confounders *a priori*. In primary analyses, we assumed that baseline indicators of TB disease severity were a result of smoking and therefore part of the pathway through which smoking could impact TB treatment outcomes (**Figure 1B**).^5–7^ However, it may be that these characteristics coincide with, but are unrelated to smoking. We therefore conducted a sensitivity analysis that additionally adjusted for these characteristics.

**Figure 1.**
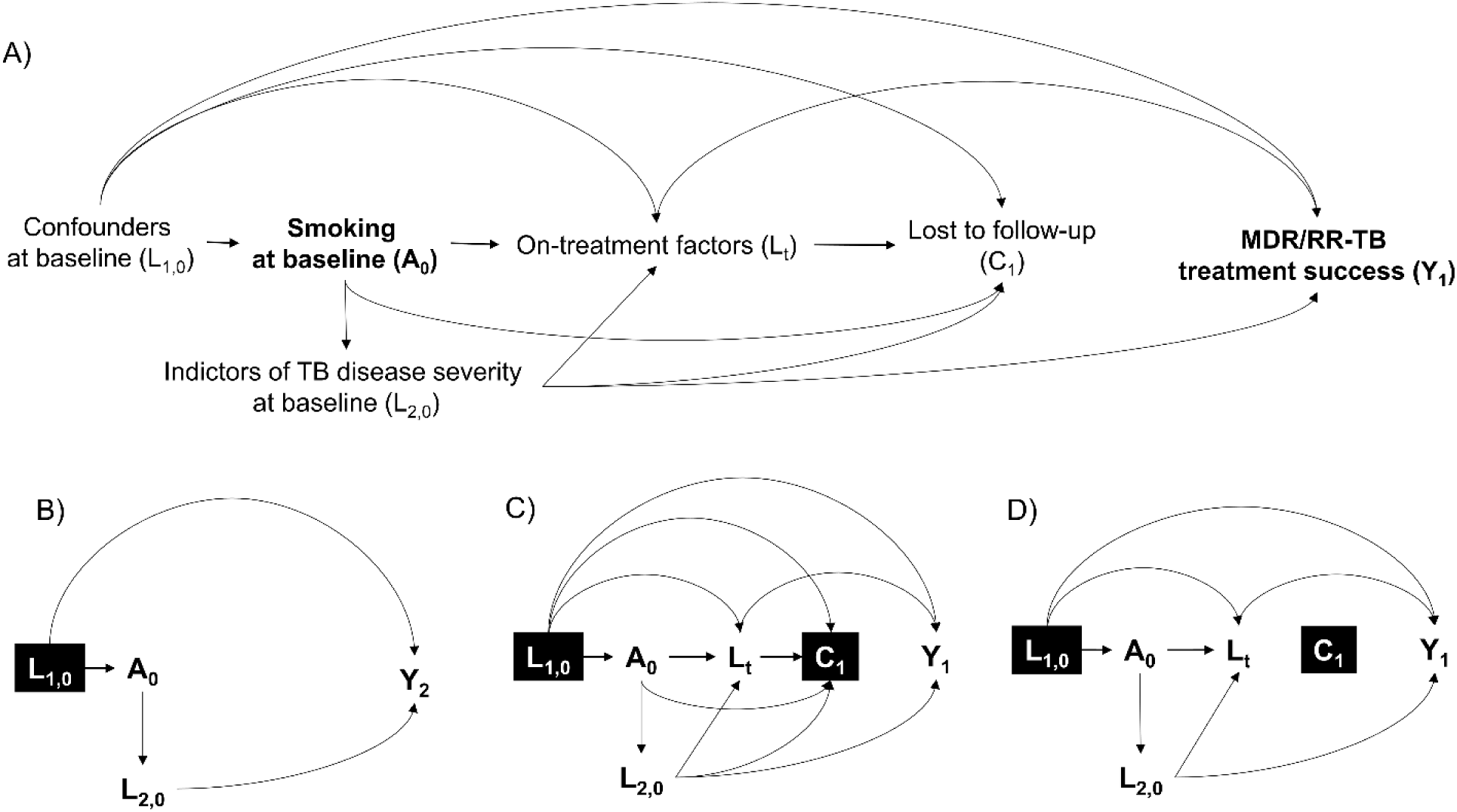
Directed acyclic graphs illustrating the relationship between cigarette smoking and MDR/RR-TB end-of-treatment outcome. L_1,0_: Baseline confounders: demographics (country, sex, age), social history (ever incarcerated, refugee/displaced person/migrant, alcohol use, drug use), and comorbidities (HIV, hepatitis C, diabetes, underweight); L_2,0_: Baseline indicators of TB disease severity (sputum smear and culture positivity, and cavitary disease); A_0_: Cigarette smoking status at enrollment; L_t_: On-treatment factors on the causal path from A_0_ to Y: time-varying indicators of TB disease severity (sputum smear and culture positivity, cavitary disease) and TB treatment adherence; C_1_: Lost to follow-up; Y_1_: Successful (cured or completed) or unsuccessful (treatment failure or death) MDR/RR-TB treatment outcome; Y_2_: Successful (cured or completed) or unsuccessful (treatment failure, death, or lost to follow-up) MDR/RR-TB treatment outcome. A) The directed acyclic graph shows the hypothesized causal relationship between cigarette smoking status at enrollment (A_0_) and a successful or unsuccessful MDR/RR-TB treatment outcome (Y_1_). Each arrow represents a one-way direct causal effect of one variable on another. B) The directed acyclic graph shows the causal relationship between A_0_ and Y_2_, adjusting for baseline confounders. Participants who were lost to follow-up were classified as not having a successful MDR/RR-TB treatment outcome. For this model, we condition on L_1,0_ (indicated by the black box), but not L_2,0_ which is expected to be on the causal path between A_0_ and Y_2_ as a mediator. As a sensitivity analysis, we explored adjusting for L_2,0_, treating it as a potential confounder (i.e., arrow changes direction between L_2,0_ and A_0_). C) The directed acyclic graph shows the causal relationship between A_0_ and Y_1_, but we additionally censor participants who were lost to follow-up (C_1_). The black box for C_1_ indicates that we are conditioning on it by limiting our analysis to participants who were not lost to follow-up. We also show additional time-varying variables (L_t_) expected to be affected by smoking and associated with loss to follow-up and the MDR/RR-TB treatment outcome. By censoring participants who were lost to follow-up, this model is unrealistic and potentially biased because it assumes that loss to follow-up was not a possible outcome during MDR/RR-TB treatment. D) The directed acyclic graph depicts the causal relationship between A_0_ and Y_1_, applying stabilized inverse probability of censoring weights to uncensored individuals (i.e., those who were not lost to follow-up) that were conditional on A_0_, L_1,0_, L_2,0_, and L_t_. Applying these weights and additionally adjusting for L_1,0_ in the final model effectively removes all of the arrows pointing to C_1_. In other words, we re-weight uncensored individuals to also account for censored individuals, creating a population where everyone is retained in treatment.

**Table 1.** Characteristics at cohort enrollment among participants included in the analysis, overall and by cigarette smoking status.

| Characteristic at cohort enrollment | n | Overall,<br>n=1786<br>n (%) | Smokes cigarettes?<br>Yes,<br>n=539<br>n (%) | No,<br>n=1247<br>n (%) |
| --- | --- | --- | --- | --- |
| *Demographics* |  |  |  |  |
| Country | 1786 |  |  |  |
| Post-Soviet country (Armenia, Belarus, Georgia, Kazakhstan, Kyrgyzstan) |  | 1056 (59.1) | 500 (92.8) | 556 (44.6) |
| Other country (Ethiopia, Indonesia, Myanmar, Pakistan, Peru, South Africa, Vietnam) |  | 730 (40.9) | 39 (7.2) | 691 (55.4) |
| Male sex | 1786 | 1136 (63.6) | 480 (89.1) | 656 (52.6) |
| Age in years, median (interquartile range; range) | 1786 | 35 (27–46; 15–83) | 41 (34–50; 17–70) | 32 (25–43; 15–83) |
| *Social history* |  |  |  |  |
| Married or living with partner | 1780 | 919 (51.6) | 259 (48.4) | 660 (53.0) |
| Employed | 1777 | 270 (15.2) | 77 (14.4) | 193 (15.5) |
| Homeless in past year | 1727 | 69 (4.0) | 24 (4.8) | 45 (3.7) |
| Ever incarcerated | 1485 | 273 (18.4) | 185 (35.4) | 88 (9.2) |
| Refugee, displaced person, or migrant | 1757 | 57 (3.2) | 33 (6.4) | 24 (1.9) |
| Drinks alcohol | 1762 | 216 (12.3) | 177 (33.5) | 39 (3.2) |
| Used non-prescribed illicit drugs and/or intravenous drugs in past year | 1718 | 93 (5.4) | 63 (12.9) | 30 (2.4) |
| *Comorbidities* |  |  |  |  |
| HIV | 1785 | 118 (6.6) | 45 (8.4) | 73 (5.9) |
| Hepatitis B virus infection | 1782 | 73 (4.1) | 27 (5.0) | 46 (3.7) |
| Hepatitis C virus infection | 1782 | 246 (13.8) | 155 (28.8) | 91 (7.3) |
| Diabetes | 1785 | 243 (13.6) | 60 (11.2) | 183 (14.7) |
| Underweight (body mass index <18.5 kg/m<sup>2</sup>) | 1777 | 647 (36.4) | 159 (29.7) | 488 (39.3) |
| *TB disease characteristics* |  |  |  |  |
| Concurrent extrapulmonary disease | 1786 | 30 (1.7) | 13 (2.4) | 17 (1.4) |
| Bilateral lung involvement | 1710 | 1137 (66.5) | 377 (71.4) | 760 (64.3) |
| Fibrosis | 1646 | 1105 (67.1) | 345 (67.9) | 760 (66.8) |
| Cavitary disease | 1676 | 1140 (68.0) | 349 (67.1) | 791 (68.4) |
| Positive sputum smear | 1683 | 946 (56.2) | 314 (61.8) | 632 (53.8) |
| Sputum smear grade (among positive) | 946 |  |  |  |
| Scanty |  | 70 (7.4) | 28 (8.9) | 42 (6.7) |
| 1+ |  | 420 (44.4) | 155 (49.4) | 265 (41.9) |
| 2+ |  | 228 (24.1) | 57 (18.2) | 171 (27.1) |
| 3+ |  | 228 (24.1) | 74 (23.6) | 154 (24.4) |
| Extensive disease (i.e., cavitary disease and smear grade 3+) | 1582 | 172 (10.9) | 53 (10.8) | 119 (10.9) |
| Positive sputum culture | 1656 | 1049 (63.4) | 350 (70.9) | 699 (60.2) |
| Known prior TB treatment with second-line drugs | 1471 | 1282 (87.2) | 405 (89.8) | 877 (86.0) |
| Fluoroquinolone resistance | 1701 | 1122 (66.0) | 358 (68.9) | 764 (64.7) |
| *Initial TB treatment regimen characteristics* |  |  |  |  |
| Number of likely effective drugs in regimen,* | 1786 | 4 (4–5; 0–8) | 4 (4–5; 0–7) | 4 (4–5; 0–8) |
| median (interquartile range; range) |  |  |  |  |
| Number of WHO Group A drugs (moxifloxacin or levofloxacin; bedaquiline; linezolid), median (interquartile range; range) | 1786 | 2 (2–3; 0–3) | 2 (2–3; 0–3) | 2 (2–3; 0–3) |
| Number of WHO Group B drugs (clofazimine; cycloserine or terizidone), median (interquartile range; range) | 1786 | 1 (1–2; 0–2) | 1 (1–2; 0–2) | 1 (1–2; 0–2) |

MDR/RR-TB, multidrug-resistant/rifampicin-resistant tuberculosis; WHO, World Health Organization.
\*A drug was considered likely effective if all reported resistance testing to that drug confirmed susceptibility, or, in the absence of drug susceptibility testing, the individual had not previously received the drug for one month or longer.

To understand if differential loss to follow-up drove the difference in treatment success by smoking status, we censored participants who were lost to follow-up and used time-varying stabilized inverse probability of censoring weights to simulate a scenario in which no one was lost to follow-up (i.e., everyone was retained in treatment; **Figure 1D**). Computation of weights included fitting a pooled logistic regression model to predict whether participants remained uncensored, conditional on smoking status; baseline demographics, social history, and comorbidities; baseline indicators of TB disease severity; and time-varying indicators of TB severity and TB medication adherence (**Supplementary Table 3**). This weighting is important because people who are lost to follow-up and those who remain in care may be different in ways that also determine the end-of-treatment outcome. We carried observations forward for time-varying variables and used the missing indicator method to account for any remaining missing data. As with the other analyses, we used marginal standardization to estimate risk differences and ratios and bootstrapping for 95% CIs.

We used SAS 9.4 (SAS Institute, Cary, NC) and R version 4.4.2 (R Core Team, 2023) for analyses.

## RESULTS

### Description of selection of the analytic study population and participant characteristics

Of 2788 endTB Observational Study participants enrolled in 17 countries, 1002 were excluded (**Figure 2**), resulting in a final analytic study population of 1786 participants in 12 countries: Armenia (93 [5.2%]), Belarus (101 [5.7%]), Ethiopia (34 [1.9%]), Georgia (208 [11.7%]), Indonesia (61 [3.4%]), Kazakhstan (641 [35.9%]), Kyrgyzstan (13 [0.7%]), Myanmar (42 [2.4%]), Pakistan (262 [14.7%]), Peru (260 [14.6%]), South Africa (39 [2.2%]), and Vietnam (32 [1.8%]). A comparison of included and excluded participants is provided in **Supplementary Table 2**; most characteristics were similar.

**Figure 2.**
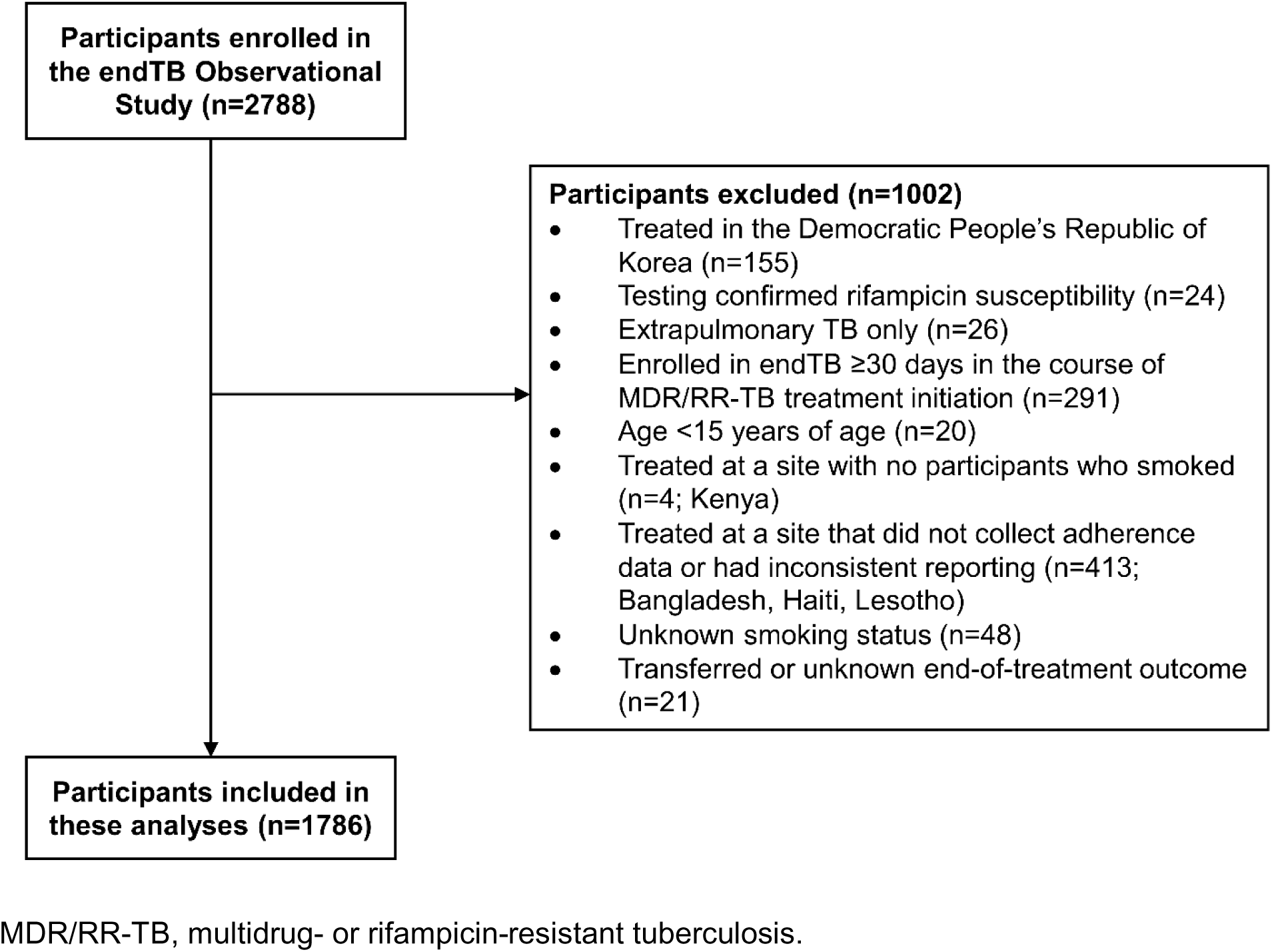
Flowchart of participants enrolled in the endTB Observational Study and their inclusion in these analyses.

Of the 1786 included participants, 539 (30.2%) reported smoking cigarettes daily. There were some notable differences in demographics, social history, comorbidities, and TB disease characteristics at enrollment by smoking status (**Table 1**). Compared with those who did not smoke, participants who smoked resided more commonly in post-Soviet countries (92.8% vs. 44.6%) and were more frequently male (89.1% vs. 52.6%), older (median age 41 years vs. 32 years), ever incarcerated (35.4% vs. 9.2%), and identified as a refugee, displaced person, or migrant (6.4% vs. 1.9%). They also more commonly drank alcohol (33.5% vs. 3.2%) and used drugs (12.9% vs. 2.4%). Regarding comorbidities, participants who smoked more often had HIV (8.4% vs. 5.9%), hepatitis B (5.0% vs. 3.7%), and hepatitis C (28.8% vs. 7.3%) and less often had diabetes (11.2% vs. 14.7%) and were underweight (29.7% vs. 39.3%) compared with those who did not smoke. Participants who smoked more often had bilateral disease (71.4% vs. 64.3%), a positive sputum smear (61.8% vs. 53.8%), and a positive sputum culture (70.9% vs. 60.2%) compared with those who did not smoke. The number of likely effective drugs in the baseline TB treatment regimen was similar by smoking status (median of 4 for both smoking categories).

### Frequencies of end-of-treatment MDR/RR-TB treatment outcomes

Overall, 1397 (78.2%) had a successful treatment outcome of cured or completed (**Table 2**). Regarding unsuccessful outcomes, 137 (7.7%) died, 97 (5.4%) had treatment failure, and 155 (8.7%) were lost to follow-up. The median (interquartile range) time from cohort enrollment to death was 5.6 (2.4, 11.5) months, and to lost to follow-up was 8.7 (5.7, 13.4) months. With regard to unsuccessful outcomes by smoking status, the frequency of death among participants who smoked was 5.0% versus 8.8% in those who did not. Compared with participants who did not smoke, the frequency of treatment failure (8.5% vs. 4.1%) and lost to follow-up (13.0% vs. 6.8%), were higher for participants who smoked.

**Table 2.** Frequencies of clinician-assigned MDR/RR-TB end-of-treatment outcomes, overall and by cigarette smoking status.

| Outcomes | Overall,<br>n=1786<br>n (%) | Smokes cigarettes? |  |
| --- | --- | --- | --- |
|  |  | Yes,<br>n=539<br>n (%) | No,<br>n=1247<br>n (%) |
| **Successful treatment** | **1397 (78.2)** | **396 (73.5)** | **1001 (80.3)** |
| Cured | 1299 (72.7) | 368 (68.3) | 931 (74.7) |
| Completed | 98 (5.5) | 28 (5.2) | 70 (5.6) |
| **Unsuccessful treatment** | **389 (21.8)** | **143 (26.5)** | **246 (19.7)** |
| Death (and causes) | 137 (7.7) | 27 (5.0) | 110 (8.8) |
| TB-related* | 64 (3.6) | 12 (2.2) | 52 (4.2) |
| Non-TB-related | 47 (2.6) | 11 (2.0) | 36 (2.9) |
| Unknown | 26 (1.5) | 4 (0.7) | 22 (1.8) |
| Treatment failure (and reasons)<sup>†</sup> | 97 (5.4) | 46 (8.5) | 51 (4.1) |
| Lack of sputum culture conversion | 32 (1.8) | 19 (3.5) | 13 (1.0) |
| Bacteriological reversion | 44 (2.5) | 15 (2.8) | 29 (2.3) |
| Adverse drug reaction | 10 (0.6) | 8 (1.5) | 2 (0.2) |
| Other/Unknown | 15 (0.8) | 7 (1.3) | 8 (0.6) |
| Lost to follow-up (and reasons)<sup>†</sup> | 155 (8.7) | 70 (13.0) | 85 (6.8) |
| Participant refusal | 71 (4.0) | 27 (5.0) | 44 (3.5) |
| Left region/country | 32 (1.8) | 21 (3.9) | 11 (0.9) |
| Family, financial, or other social problems | 35 (2.0) | 12 (2.2) | 23 (1.8) |
| Substance abuse | 12 (0.7) | 5 (0.6) | 7 (0.6) |
| Other/Unknown | 36 (2.0) | 14 (2.6) | 22 (1.8) |

\*TB was known to be an immediate or contributing cause of death, or death was related to surgery/treatment of TB.
<sup>†</sup>Participants may have more than one clinician-assigned reason for treatment failure (with the exception of simultaneous lack of sputum culture conversion and bacteriological reversion) or loss to follow-up.

### Effect of smoking on MDR/RR-TB treatment success and eliminating loss to follow-up

In the unadjusted model (**Table 3**), the risk of successful treatment outcome was 6.8 percentage points lower among participants who smoked (73.5%) versus those who did not (80.3%; 95% CI for risk difference: -11.1, -2.6) with a risk ratio of 0.92 (95% CI: 0.86, 0.97). When adjusting for baseline confounders related to demographics, social history, and comorbidities, the risk difference was -5.2 percentage points (95% CI: -14.1, 3.2) and the risk ratio was 0.93 (95% CI: 0.82, 1.04). In the sensitivity analysis that also adjusted for baseline indicators of TB disease severity, the risk difference was -4.4 percentage points (95% CI: -12.7, 3.2) and the risk ratio was 0.94 (95% CI: 0.84, 1.04).

**Table 3.** Effect of smoking status on a successful MDR/RR-TB end-of-treatment outcome and impact of intervening on loss to follow-up.

| Outcome and analysis* | Risk, percentage |  | Risk difference, percentage points (95% CI) | Risk ratio (95% CI) |
| --- | --- | --- | --- | --- |
|  | Participants who smoked | Participants who did not smoke |  |  |
| Successful treatment (with loss to follow-up classified as unsuccessful treatment) |  |  |  |  |
| Unadjusted | 73.5 | 80.3 | -6.8 (-11.1, -2.6) | 0.92 (0.86, 0.97) |
| Adjusted for baseline confounders<sup>†</sup> | 73.8 | 79.0 | -5.2 (-14.1, 3.2) | 0.93 (0.82, 1.04) |
| Adjusted for baseline confounders<sup>†</sup> and baseline indicators of TB disease severity<sup>‡</sup> (sensitivity analysis) | 74.5 | 78.9 | -4.4 (-12.7, 3.2) | 0.94 (0.84, 1.04) |
| Successful treatment (with everyone retained in treatment) |  |  |  |  |
| IP weighted and adjusted for baseline confounders<sup>†</sup> | 84.0 | 85.9 | -1.9 (-11.1, 4.7) | 0.98 (0.87, 1.05) |

CI, confidence interval; IP, inverse probability; TB, tuberculosis.
\*Generalized linear models with logit links were used to estimate risks separately by smoking status and risk differences and ratios were computed by standardizing mean predicted risks of the outcome. The bias-corrected and accelerated bootstrap with 1000 resamples was used to obtain 95% confidence intervals for the risk differences and ratios. The missing indicator method used to handle missing data on baseline confounders except for HIV status (n=1 [0.1%]), hepatitis C (n=4 [0.2%]), and diabetes (n=1 [0.1%]) for which we assumed the condition to be absent because very few missing observations precluded use of missing indicators and these were low prevalence conditions.
<sup>†</sup>Demographics (country, sex, age), social history (ever incarcerated, refugee/displaced person/migrant, drinks alcohol, drug use), and comorbidities (HIV, hepatitis C, diabetes, underweight).
<sup>‡</sup>Sputum and culture positivity, and presence of cavitary disease.

When eliminating loss to follow-up, the risk of successful treatment was (84.0%) among participants who smoked compared with (85.9%) in those who did not (risk difference -1.9 percentage points, 95% CI: -11.1, 4.7; risk ratio: 0.98, 95% CI: 0.87, 1.05).

## DISCUSSION

Cigarette smoking was common at the start of MDR/RR-TB treatment, particularly in post-Soviet countries, and was frequently accompanied by a complex social history, including past incarceration and substance use, and infectious comorbidities, especially hepatitis C. We found that people who smoked had a lower frequency of MDR/RR-TB treatment success than those who did not smoke. Simulating elimination of loss to follow-up (i.e., ensuring everyone completed treatment) attenuated the difference in treatment success by smoking status. This finding suggests that smoking may negatively impact MDR/RR-TB treatment outcomes through complex adherence-related pathways leading to loss to follow-up. However, since some difference by smoking status remained, other pathways like the direct biological pathways leading to treatment failure are relevant.

In the model adjusted for baseline confounders, the risk difference of treatment success by smoking status was -5.2 percentage points, which is clinically meaningful and similar to that observed in other studies. For example, in a large prospective cohort from Hong Kong with 16,345 people who had drug-susceptible TB, the risk difference in treatment success by smoking status (current vs. never) was about -3.3 percentage points.^7^ When we adjusted for confounders, the CI was less precise than for the unadjusted estimate and included the null value of 1. However, since we estimated causal effects, we focus our interpretation on magnitude and precision rather than solely statistical significance (i.e., p<0.05).^32,33^ Despite less precision, the adjusted risk difference was similar in magnitude to the unadjusted risk difference and thus baseline confounding did not appear to be a major driver of the observed association. Selection of confounders requires a clear understanding of temporality relative to the exposure, which may not always be known. For example, more advanced TB disease may result from smoking through direct biological effects, but may also coincide with smoking if people who smoke have delays in diagnosis (e.g., if they are less likely to identify TB symptoms as indicative of illness). In our sensitivity analysis, adjusting for sputum smear and culture positivity, and presence of cavitary disease slightly attenuated the risk difference, but did not lead to a different conclusion. Because many social factors and medical conditions can influence how TB presents, we recommend conducting sensitivity analyses to assess how assumptions about whether a variable is a confounder or part of the causal pathway may affect the results.

To understand the extent to which adverse outcomes among people who smoked were explained by treatment lapses versus the biological effects of smoking, we estimated the effect of smoking in the absence of loss to follow-up. In practical terms, this analysis was analogous to implementing an intervention that equalized retention in treatment by smoking status. We found that the risk difference in treatment success by smoking status decreased from -5.2 to -1.9 percentage points, supporting that complex adherence-related pathways leading to loss to follow-up were a driver of worse treatment outcomes among people who smoked. Although this finding may seem intuitive, eliminating loss to follow-up would not be expected reduce the disparity among people who smoked if, for example, poor clinical evolution contributed to early treatment discontinuation. If this were true, eliminating loss to follow-up could result in retention of individuals at increased risk of treatment failure. Our analytic approach accounted for this potential scenario by including time-varying factors associated with loss to follow-up and unfavorable treatment outcomes (i.e., measures of TB disease severity and TB medication adherence) in our inverse probability of censoring weights. The attenuated risk difference when eliminating loss to follow-up suggests that interventions that facilitate retention in treatment, particularly among people who smoke, could reduce the observed difference in TB treatment outcomes by smoking status. The reasons why people disengage from MDR/RR-TB treatment are complex,^18,34,35^ but evidence supports interventions providing psychosocial support throughout TB treatment, such as through counseling sessions and home visits by healthcare workers, as one possible way.^36^ Adding conditional cash transfers to psychosocial support interventions may also address economic factors that contribute to loss to follow-up.^37^ Although such interventions might be broadly beneficial for people undergoing TB treatment, further research is needed to understand how reasons for loss to follow-up might differ for specific groups (e.g., people who smoke) to optimize and better target interventions.

The remaining difference in TB treatment outcomes after eliminating loss to follow-up could plausibly be attributed to random variability, but most likely to a direct biological effect of smoking on TB treatment failure. Evidence from drug-susceptible TB supports that smoking cessation after a TB diagnosis and continuing not to smoke during treatment may increase treatment success and lower recurrence compared with those who continue to smoke.^38,39^ Recognizing the intersections in TB and lung health, the World Health Organization recommends an integrated approach to care,^40^ for example, implementing guideline recommendations on tobacco cessation^41^ into the delivery of MDR/RR-TB treatment. Multiple studies in various high burden countries have demonstrated the feasibility of integrating smoking cessation into TB treatment and care programs.^42–44^ Implementation research would be beneficial to optimize integration of services and should consider the multiple issues that face people with TB, not limited to complex social and economic contexts and comorbidities related to both physical and mental health.

Aside from leveraging a large, high-quality observational cohort of people with MDR/RR-TB with extensive data collection, the major strength of this analysis lies in its methodology.

Specifically, we applied a causal inference framework when designing the analysis, including the use of directed acyclic graphs to transparently guide analysis decisions and weighting to appropriately account for baseline and on-treatment factors that predicted loss to follow-up. Such methodology is novel in the field of TB and tobacco and its application in this analysis contributes to existing knowledge. Our findings should also be interpreted in the context of limitations. The definition of smoking status is subject to potential misclassification. For example, some participants who smoked might have chosen to report that they did not smoke and since smoking was only assessed at enrollment, we do not know if individuals quit smoking during treatment. We would expect that each of these sources of potential misclassification would diminish differences in TB treatment outcomes by smoking status, biasing toward the null. Despite analyzing a large cohort, we had limited precision in some analyses. However, since we aimed to estimate causal effects, our interpretation focused on the magnitude and precision of these effects instead, as recommended by causal inference experts and the American Statistical Association.^32,33^ To increase confidence in our findings, we would recommend replication in other cohorts. Most people who smoked were in post-Soviet countries, so although we leveraged a multicountry cohort, generalizability may still be limited. Despite our best efforts to identify and control for potential confounders, residual and unmeasured confounding (e.g., due to mental health morbidity), might have resulted in bias.

In conclusion, people who smoked had a lower frequency of MDR/RR-TB treatment success than those who did not smoke. Eliminating loss to follow-up substantially reduced, but did not eliminate, the difference in treatment outcomes experienced by people who smoked. This finding suggests that complex pathways related to retention in treatment were the driver of this disparity, although other pathways, such as those related to the direct biological effects of smoking, may also contribute. Implementing interventions to address causes of loss to follow-up, e.g., counseling and home visits, and financial support, and integrating smoking cessation services could improve MDR/RR-TB treatment outcomes among people who smoke.

## Data Availability

All data produced in the present study are available upon reasonable request to the authors. endTB Observational Study data are also available upon request: https://endtb.org/access-endtb-data-through-edsi

## Ethics approval

The endTB Observational Study protocol was approved by ethics committees for each consortium partner (Partners Healthcare: #2015P001669; IRD: #IRD_IRB_2015_08_001; MSF: #1550) and in each country.

## SUPPLEMENTARY TABLES

**Supplementary Table 1.** Rationale for each exclusion criterion for the analytic study population.

| Exclusion criterion | Rationale |
| --- | --- |
| Treated in the Democratic People's Republic of Korea | Compared with other sites, the Democratic People's Republic of Korea had major differences in clinical protocols (e.g., no HIV testing) and regimens, including shortened treatments. For these reasons, other analyses of endTB Observational Study data have typically excluded this country. |
| Had confirmed rifampicin-susceptible TB | Our analysis was focused on people with MDR/RR-TB. Some participants, particularly when culture-based methods for resistance were used, may have experienced a delay between cohort enrollment and obtaining rifampicin resistance results. |
| Had only extrapulmonary TB | The biologic pathways in which smoking affects TB treatment outcomes for extrapulmonary TB may differ from those for pulmonary TB (lung damage and impaired pulmonary immune response). Some variables used in our analyses are not applicable to people without pulmonary TB (i.e., sputum smear and culture positivity, presence of cavitory disease). |
| Enrolled in the endTB Observational Study more than one month after MDR/RR-TB treatment initiation | Some participants in the endTB Observational Study were enrolled later in the course of MDR/RR-TB treatment and their time of observation would not align with that of the majority of participants who started treatment around the time of cohort enrollment. This was an important consideration because of how the exposure was ascertained (smoking at baseline, i.e., start of MDR/RR-TB treatment vs. later during treatment) and our longitudinal analyses, which included time-varying variables. |
| Were under 15 years of age | Adults and older adolescents ($\geq 15$ years of age) comprise the vast majority of people who smoke cigarettes daily and, therefore, are our target population. Furthermore, reasons for lost to follow-up for younger participants (0–14 years of age) are expected to be different from those for older participants and likely involve their parents or caregivers. |
| Treated at a site with no participants who reported smoking | Recognizing that there are both measured and unmeasured differences in participant characteristics among sites, we sought to include sites that had at least some variation in the exposure (smoking). |
| Treated at a site that did not collect adherence data or had inconsistent reporting | Since we expected TB treatment adherence to be strongly predictive of loss to follow-up, it was used in our computation of inverse probability of censoring weights. We would not be able to compute these weights for participants at sites that did not collect adherence data, and we would not be able to accurately compute these weights for participants at sites with inconsistent reporting. |
| Transferred out or had an unknown end-of-treatment outcome | This small number of participants would be censored in our analyses since their MDR/RR-TB outcome was unknown. Their exclusion from the analytic sample is unlikely to influence our results. |

**Supplementary Table 2.** Selected characteristics of endTB Observational Study participants stratified by inclusion and exclusion into our analytic study population.

| Characteristic at cohort enrollment | n | All included,*<br>n=1786<br>n (%) | n | Excluded, but at same sites as included,†<br>n=260<br>n (%) |
| --- | --- | --- | --- | --- |
| Post-Soviet country | 1786 | 1056 (59.1) | 260 | 141 (54.2) |
| Male sex | 1786 | 1136 (63.6) | 260 | 159 (61.2) |
| Married or living with partner | 1780 | 919 (51.6) | 255 | 111 (43.5) |
| Employed | 1777 | 270 (15.2) | 251 | 39 (15.5) |
| Homeless in past year | 1727 | 69 (4.0) | 241 | 8 (3.3) |
| Ever incarcerated | 1485 | 273 (18.4) | 203 | 35 (17.2) |
| Refugee, displaced person, or migrant | 1757 | 57 (3.2) | 239 | 9 (3.8) |
| Drinks alcohol | 1762 | 216 (12.3) | 234 | 25 (10.7) |
| Used non-prescribed illicit drugs and/or intravenous drugs in past year | 1718 | 93 (5.4) | 221 | 15 (6.8) |
| HIV | 1785 | 118 (6.6) | 260 | 31 (11.9) |
| Hepatitis B virus infection | 1782 | 73 (4.1) | 258 | 16 (6.2) |
| Hepatitis C virus infection | 1782 | 246 (13.8) | 260 | 29 (11.2) |
| Diabetes | 1785 | 243 (13.6) | 259 | 30 (11.6) |
| Bilateral pulmonary disease | 1710 | 1137 (66.5) | 241 | 138 (57.3) |
| Lung fibrosis | 1646 | 1105 (67.1) | 224 | 130 (58.0) |

\*In **Table 1**. †A total of 847 participants were excluded, including all participants at four sites: Bangladesh (n=279), Haiti (n=37), Kenya (n=7), Lesotho (n=264). Because of expected differences by sites, these 587 participants were not included for this comparison.

**Supplementary Table 3.** Specification of stabilized time-varying inverse probability of censoring weights.

| Specification | Mean<br>(SD) | Min,<br>max |
| --- | --- | --- |
| **Numerator:** Time;* quadratic time; smoking status ($A_0$); and baseline demographics, social history, and comorbidities ($L_{1,0}$).<sup>†</sup><br>**Denominator:** Time;* quadratic time; smoking status ($A_0$); and baseline demographics, social history, and comorbidities ($L_{1,0}$);<sup>†</sup> baseline indicators of TB disease severity ($L_{2,0}$),<sup>†</sup> time-varying measures of TB diseases severity (smear positivity, culture positivity, and cavitary disease) and TB treatment adherence ($L_t$).<sup>‡</sup> | 1.00<br>(0.11) | 0.58,<br>2.34 |

SD, standard deviation.
\*Time defined as week of treatment.
<sup>†</sup>As with the regression analyses, a missing category was included in the variable and separate variable to indicate missingness was added.
<sup>‡</sup>For time-varying variables, observations were carried forward in time until measured again or the end of treatment. For any missing data, a missing category was included in the variable and separate variable to indicate missingness was added.

